# The burden of neurogenic orthostatic hypotension in patients with multiple system atrophy – a real-world study

**DOI:** 10.64898/2026.04.20.26351214

**Authors:** Matthew J. Kmiecik, Leah O’Brien, Molly Szpyhulsky, Valeria Iodice, Roy Freeman, Jens Jordan, Italo Biaggioni, Horacio Kaufmann, Ross Vickery, Aine Miller, Emma Saunders, Emma Rushton, Lesley Valle, Lucy Norcliffe-Kaufmann

## Abstract

**Background:** Although neurogenic orthostatic hypotension (nOH) is a common and debilitating feature of multiple system atrophy (MSA), little is known about the burden of symptoms in the real world.

**Objectives:** To design and conduct a cross-sectional community-based research survey targeting patients with MSA, with and without nOH.

**Methods:** We recruited patients with MSA to complete an anonymous online survey covering three core themes: 1) timely diagnosis, 2) nOH pharmacotherapy and refractory symptoms, and 3) confidence in physician knowledge. Responses were grouped by pre-specified diagnostic certainty levels. Relationships between symptoms, function, and pharmacotherapy were assessed using univariate and multivariate methods.

**Results:** We analyzed 259 respondents with a self-reported diagnosis of MSA (age: *M*=64.38, *SD*=8.09 years; 44% female). In total, 42% also had a diagnosis nOH; 40% had symptoms highly suspicious of nOH, but no diagnosis; and 21% reported having never had their blood pressure measured in the standing position at a clinical visit. Treatment with a pressor agent was independently associated with the presence of other symptoms of autonomic failure. Each additional nOH symptom reported increased the odds of requiring pharmacotherapy by 18%. Yet, despite anti-hypotensive medication use, 97% of patients reported limitations in their ability to bathe, cook, or arise from a chair/bed with 76% needing caregiver support for refractory nOH symptoms.

**Conclusions:** This cross-sectional representative sample shows nOH is underrecognized and undertreated in MSA patients, leading to substantial functional limitations. It is our hope that these findings are leveraged for planning future trials and advocating for better treatments.

---

Multiple system atrophy (MSA) is a rare disease in which 80% of patients will develop neurogenic orthostatic hypotension (nOH) in their lifetime [1, 2]. This can occur either as an initial symptom of neurodegeneration in the prodromal stage or after the threshold for clinical diagnosis is met and the disease progresses [3, 4]. Pathologically, nOH in a patient with MSA arises from synuclein-associated degeneration of the central autonomic network with relative sparing of the peripheral autonomic neurons [5]. Despite normal circulating levels of the vasoconstrictor neurotransmitter norepinephrine, this central autonomic lesion prevents the regulation of blood pressure (BP) in response to gravitational (orthostatic) stress, leading to hypotension when upright [6, 7]. The cardinal symptoms of nOH develop due to hypoperfusion of the organs, when blood supply falls below a critical autoregulatory limit, causing tissue ischemia [8, 9]. Global cerebral ischemia stemming from severe nOH can result in syncope, frequent falls, and cognitive impairment [10, 11]. Over the long-term, chronic nOH predicts poor prognosis and early death from all-cause mortality [12].

Despite nOH being a common feature of MSA, it remains unclear how the burden of symptoms impacts activities of daily living and contributes to disability and dependency. No dedicated study has looked at symptoms of nOH in the real world. Most of what is known comes from small in-clinic datasets that are geographically restricted to single center studies or small cohort-based observational trials that come from autonomic laboratories [13–15]; both are prone to referral bias. It appears that only a weak relationship exists between BP and symptoms, with no discrete cut off, and patients being asymptomatic or mildly symptomatic despite substantial BP falls [16]. Consequently, the perception of nOH symptoms, the limitations they impose on life, and influence of background disease make it unclear which patients to treat with anti-hypotensive medications. Moreover, the sparsity of data becomes a challenge when planning clinical trials with subjective symptom measures as endpoints. Without understanding fluctuations in nOH, symptom variability, and interactions with comorbidities, it remains challenging to plan sample sizes, enrollment criteria, and strategies to minimize noise to measure treatment effects. It remains unclear whether nOH symptoms are a feasible study endpoint as sample size is limited by the low prevalence and variable clinical expression of MSA.

Well-designed and well-executed surveys can provide critical insights into rare diseases in diverse representative target populations [17]. Drawing on this approach, we developed an online survey instrument with the specific research goal of documenting the unmet needs in 1) timely diagnosis, 2) pharmacotherapy, and 3) physician-knowledge of MSA and nOH from patients in the United Kingdom (UK) and United States (US). To uncover the complex relationship between function, nOH, and background disease, we applied both univariate approaches and a multivariate principal component analysis to interpret the dataset.

## Methods

### Survey design and development

We conducted a literature review and adapted existing instruments where possible [18–20] to outline an anonymous survey draft. We convened a committee composed of six experts with backgrounds in movement disorders and autonomic failure, and two specialist MSA nurses, biostatisticians, and patient representatives to identify core themes. We outlined a series of 28 patient-facing English-language questions and answers with branching logic (see Supplemental Material). As patients were not familiar with the term “neurogenic OH”, throughout the survey, we used the phrase “orthostatic or postural hypotension due to a fall in blood pressure when standing or walking” to avoid confusion. Patient leaflets and frequently asked questions documents were custom designed to explain the study objectives, data use, and funding sources (see Supplemental Material).

The cross-sectional survey was programmed using the SurveyMonkey platform in collaboration with psychometricians. An online landing page was built with a web-based consent process and MSA accessibility features to mitigate non-responses. Prior to launch, the survey design was beta-tested for plausibility and completion rates in four MSA patients. Survey responses were completely anonymous and no identifying information was collected. The study protocol and recruitment materials were reviewed by two external ethics committees (US: Advarra and UK: Health Research Authority Research Ethics Committee), and exemptions were granted in both countries. Access to response data was restricted to authorized study personnel via password-protected accounts, preventing unauthorized access.

### Population and Recruitment

The survey targeted patients with a self-reported diagnosis of MSA, at least 18 years old, in the US and UK via convenience sampling. Recruitment materials were designed around the theme of patients being able to contribute to research from their own homes. Links to the landing page were distributed from November 2025 to February 2026 through emails from patient organizations (MSA Trust UK, Defeat MSA, and Mission MSA), social media posts, community representatives, and support group events (UK only). Individualized links were created for each organization to identify the most successful recruitment initiatives.

### Measures

The survey had four core domains: (i) *Diagnostic journey*: including age at first symptom(s), physician encounters, age at diagnosis, diagnosing physician specialty, current symptoms, and extent of motor impairment; (ii) *Symptoms of nOH*: presence/absence of nOH symptoms, relationship with posture, functional limitations imposed by nOH, symptom severity, and need for caregiver support related to nOH; (iii) *nOH pharmacotherapy, complications, and refractory symptoms*: including current lifestyle/pharmacological interventions, presence of supine hypertension, and aspirations for future treatments; (iv) *Awareness of nOH*: including physician knowledge, screening of orthostatic vitals during clinic visits, and accessibility to home BP measuring devices. Dichotomous questions included yes/no and a “not sure” option. General questions included multiple-choice options. Additional open-ended questions allowed respondents to report the presence of symptoms in advanced-stage disease for those unable to stand. To maximize the number of diagnosed respondents, free-text typed responses that aligned available option choices were assigned to intended response.

### Data analysis

The protocol included a data analysis plan with pre-specified subgroups (see Figure 1). To enrich the cohort for a higher level of diagnostic certainty, those ≥80 years of age at MSA diagnosis, who did not report at least one motor deficit and at least one autonomic symptom, and did not receive a diagnosis of MSA from a specialist neurologist, sleep medicine doctor, or autonomic physician—were excluded from analysis.

**Figure 1.**
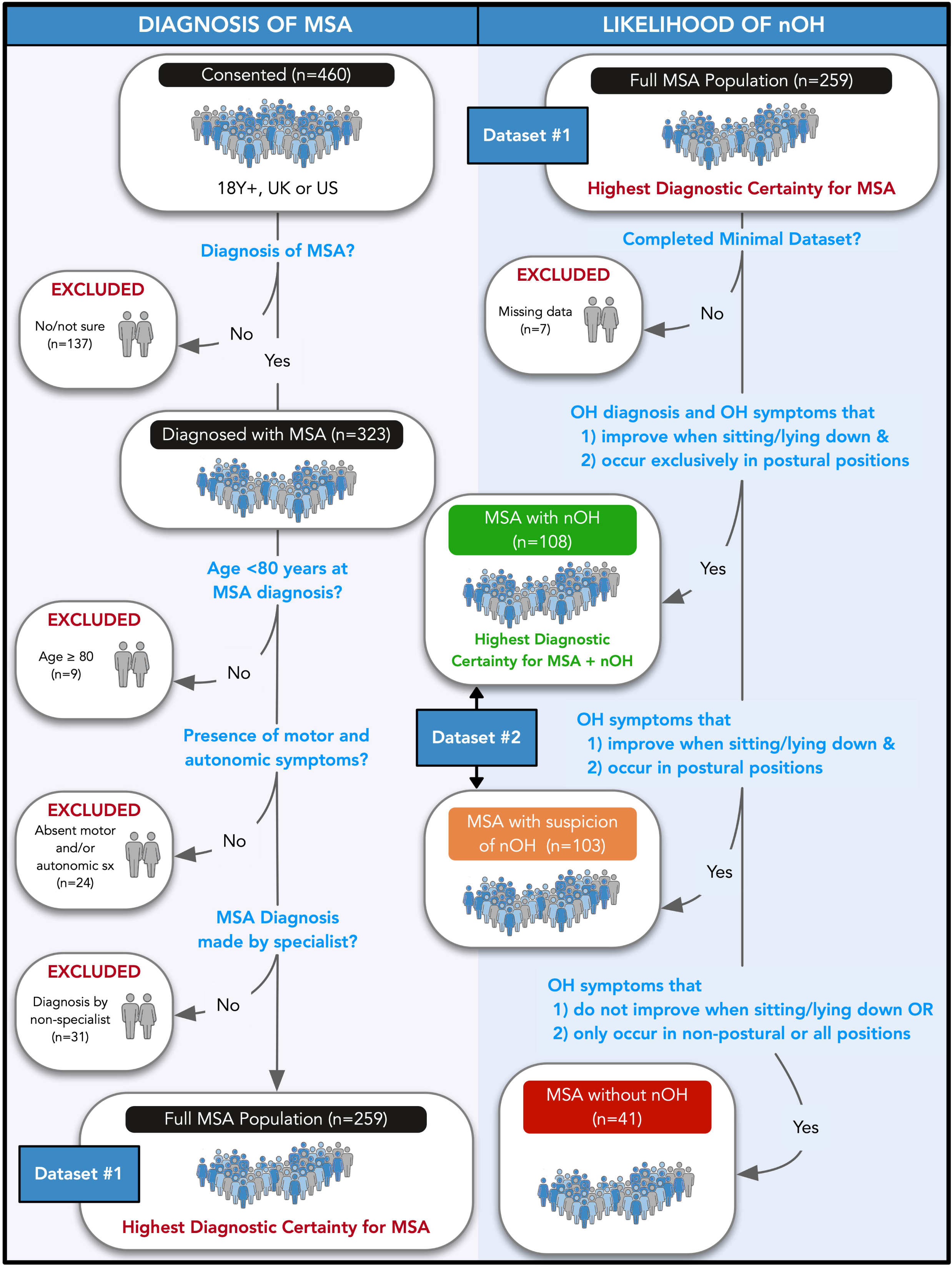
Strategic subgroup assignment flowchart depicting diagnostic certainty criteria for MSA and nOH. From the Full MSA Population, participants were further classified into MSA with: nOH, suspicion of nOH, and without nOH.

Our analysis aims were two-fold: 1) to provide descriptive statistics across pre-defined groups of MSA and nOH to define unmet need, 2) to understand the relationship between MSA/nOH symptoms and their associated functional burden, and 3) to identify a subpopulation of patients with more severe nOH symptoms in need of treatment with anti-hypotensive medications.

In MSA patients with nOH and suspicion of nOH (dataset #2, see Figure 1), the following analyses were performed. The relationship between nOH symptoms and functional burden was estimated using a Pearson correlation with a bootstrapped 95% confidence interval (CI). We used logistic regression to estimate the association between anti-hypotensive medication use and the number of nOH symptoms, adjusting for age and sex. We established cut-points for the number of nOH symptoms that increased the relative odds of pharmacotherapy with predicted probability curves (holding age constant at the sample mean) and by maximizing the Youden index (J = sensitivity + specificity - 1) [21].

To understand the interactions between motor deficits, autonomic impairment, cognition/mood, and nOH symptoms and how they contribute to functional burden and medication use, we performed multiple correspondence analysis (MCA), a variant of principal component analysis using dummy-coded categorical variables [22]. We performed permutation testing for principal components (PCs) and bootstrapping for stability/significance of loadings with 2,000 iterations. Bootstrap ratios were calculated by dividing the observed loadings by their standard deviations derived from bootstrapped supplementary projections [23]. We interpreted the PCs by examining the scree plot with permutation testing results, bootstrap ratios, and geometrically plotted loadings. We regressed functional burden and nOH-related BP medication usage on significant PCs, using linear and logistic regression, respectively. Age and sex served as covariates. PCs and age were z-scored prior to modeling (*M*=0, *SD*=1). Participants with missing data on any variable included in each model were excluded from that analysis (listwise deletion) and no sample weighting was performed.

Data wrangling, statistical analysis, and data visualization was performed in R (v.4.4.1; [24]) using the *tidyverse* [25], *correlation* [26], *patchwork* [27], and *ExPosition* [23] packages. Data are available on Open Science Framework (link to be inserted here upon publication).

## Results

### Respondents and diagnostic journey

Table 1 shows the key demographics and clinical characteristics of the cohort and data sets. Because participants were recruited through social media channels and patient advocacy networks, a formal response rate could not be calculated. Of 460 individuals who initiated the survey, 325 completed it. The full MSA population included 259 self-reported cases (mean±SD age 64.38±8.09 years, 56% male), with compatible symptoms, < 80 years of age, who completed the survey with a median time of 16 minutes. Mean age at first symptom was 58.51±8.15 years and mean age at MSA diagnosis was 61.64±7.76 years. On average, diagnostic delay was 3.13 years. Initially, 70% of patients were first told they had a different diagnosis to MSA. The most common misdiagnoses were Parkinson disease (47.5%), cerebellar ataxia (13.3%), and dysautonomia or pure autonomic failure (5.0%), with a small percentage of patients (3.9%) having first been told they had a psychological disorder. On average, patients sought care from 4 different doctors for symptoms later found to be related to MSA. Most patients eventually received an MSA diagnosis from a neurologist (93.4%), with 74.0% recalling being first diagnosed by movement disorder specialist. In a small number of cases, the first diagnosis was made by an autonomic specialist (4.6%) or sleep medicine physician (1.9%).

**Table 1.** Demographics and clinical characteristics of the MSA cohort and subgroups according to diagnostic certainty of nOH.

| Parameter | Full MSA Population | MSA with nOH | MSA with Suspicion of nOH | MSA Without nOH |
| --- | --- | --- | --- | --- |
| **Diagnostic Journey** |  |  |  |  |
| n (% of Full MSA Population) | 259 (100%) | 108 (41.7%) | 103 (39.77%) | 41 (15.83%) |
| Sex (Female) | 114 (44.02%) | 52 (48.15%) | 39 (37.86%) | 19 (46.34%) |
| Current Age (y) | 64.38 (0.5) | 65.28 (0.81) | 63.86 (0.77) | 62.41 (1.23) |
| Age at First Symptom (y) | 58.51 (0.51) | 59.22 (0.8) | 58.02 (0.79) | 56.9 (1.31) |
| Physicians Encountered Prior to Diagnosis | 4.02 (0.22) | 4.14 (0.3) | 4.16 (0.38) | 3.59 (0.53) |
| Age at MSA Diagnosis (y) | 61.64 (0.48) | 62.31 (0.76) | 61.41 (0.74) | 59.51 (1.2) |
| MSA Disease Duration (y) | 2.74 (0.16) | 2.96 (0.31) | 2.46 (0.2) | 2.9 (0.28) |
| Different Initial Diagnosis | 180 (69.77%) | 74 (68.52%) | 70 (68.63%) | 32 (78.05%) |
| **Current Symptoms** |  |  |  |  |
| Clumsiness or Tremor | 242 (93.44%) | 99 (91.67%) | 97 (94.17%) | 40 (97.56%) |
| Falls | 182 (70.27%) | 80 (74.07%) | 65 (63.11%) | 33 (80.49%) |
| Dysarthria | 226 (87.26%) | 93 (86.11%) | 88 (85.44%) | 39 (95.12%) |
| Dysphagia | 137 (52.9%) | 52 (48.15%) | 56 (54.37%) | 26 (63.41%) |
| Orthostatic Lightheadedness | 190 (73.36%) | 94 (87.04%) | 72 (69.9%) | 20 (48.78%) |
| Hypohidrosis | 120 (46.33%) | 54 (50%) | 48 (46.6%) | 14 (34.15%) |
| Constipation or Incontinence | 192 (74.13%) | 86 (79.63%) | 73 (70.87%) | 27 (65.85%) |
| Urinary Retention | 104 (40.15%) | 43 (39.81%) | 42 (40.78%) | 16 (39.02%) |
| Nocturia | 132 (50.97%) | 56 (51.85%) | 50 (48.54%) | 24 (58.54%) |
| Urinary Catheter Use | 92 (35.52%) | 36 (33.33%) | 33 (32.04%) | 20 (48.78%) |
| Frequent UTIs | 83 (32.05%) | 35 (32.41%) | 31 (30.1%) | 14 (34.15%) |
| Orthopedic Deformities of Neck or Spine | 102 (39.38%) | 42 (38.89%) | 42 (40.78%) | 15 (36.59%) |
| Stridor | 93 (35.91%) | 36 (33.33%) | 36 (34.95%) | 16 (39.02%) |
| Dream Re-Enactment Behaviors | 175 (67.83%) | 73 (67.59%) | 69 (66.99%) | 28 (68.29%) |
| Depression | 128 (49.61%) | 56 (51.85%) | 50 (48.54%) | 21 (51.22%) |
| Anxiety | 133 (51.55%) | 56 (51.85%) | 58 (56.31%) | 18 (43.9%) |
| Pseudo-bulbar Affect | 100 (38.76%) | 40 (37.04%) | 38 (36.89%) | 18 (43.9%) |
| **Degree of Disability** |  |  |  |  |
| Requires No Assistance or Device | 42 (16.28%) | 18 (16.67%) | 19 (18.45%) | 4 (9.76%) |
| Use of Walker or Cane | 76 (29.46%) | 38 (35.19%) | 32 (31.07%) | 6 (14.63%) |
| Occasional Use of Wheelchair | 50 (19.38%) | 22 (20.37%) | 20 (19.42%) | 6 (14.63%) |
| Frequent Use of Wheelchair | 67 (25.97%) | 23 (21.3%) | 24 (23.3%) | 17 (41.46%) |
| In Bed Most of Time | 23 (8.91%) | 7 (6.48%) | 8 (7.77%) | 8 (19.51%) |
| **nOH Specific Symptoms When Upright** |  |  |  |  |
| Frequent Fainting | 61 (24.02%) | 32 (30.19%) | 19 (18.81%) | 8 (19.51%) |
| Lightheadedness/Dizziness | 183 (72.05%) | 95 (87.96%) | 72 (69.9%) | 16 (39.02%) |
| Fatigue/Tiredness | 196 (77.17%) | 86 (79.63%) | 80 (77.67%) | 29 (70.73%) |
| Weakness | 189 (74.41%) | 83 (76.85%) | 79 (76.7%) | 25 (60.98%) |
| Coat Hanger Pain | 129 (50.79%) | 55 (50.93%) | 55 (53.4%) | 18 (43.9%) |
| Visual Problems | 119 (46.85%) | 53 (49.07%) | 48 (46.6%) | 18 (43.9%) |
| Concentration/Attention Difficulties | 100 (39.37%) | 41 (37.96%) | 42 (40.78%) | 17 (41.46%) |
| Difficulties Responding to Questions | 77 (30.31%) | 32 (29.63%) | 33 (32.04%) | 12 (29.27%) |
| Shortness of Breath | 79 (31.1%) | 28 (25.93%) | 32 (31.07%) | 18 (43.9%) |
| Chest Pain/Pressure | 17 (6.69%) | 6 (5.56%) | 7 (6.8%) | 4 (9.76%) |

| Parameter | Full MSA<br>Population | MSA with<br>nOH | MSA with<br>Suspicion of<br>nOH | MSA<br>Without<br>nOH |
| --- | --- | --- | --- | --- |

*Note.* Continuous parameters are presented Mean (SEM); categorical parameters are presented n (%); the 'n' row percentage represents each subgroup's proportion of the Full MSA Population; all other row percentages are calculated from valid (non-missing) responses within each group; nOH=neurogenic orthostatic hypotension; MSA=multiple system atrophy; UTI=urinary tract infection.

### Symptoms of MSA

In the full MSA sample, mean time since diagnosis of MSA was 2.74±2.57 years. The most frequently reported motor symptoms were clumsiness, tremor or jerking (93.4%), dysarthria (slurred/soft speech: 87.3%), gait imbalance (falls: 70.3%), and dysphagia (coughing or choking: 52.9%). All respondents reported at least one non-motor symptom with the most common being urinary dysfunction (88.0%). Physical capabilities varied from being able to walk with no assistance (16.3%), use of a cane or walker (29.5%), intermittent use of a wheelchair (19.4%), being wheelchair bound (26.0%), to being bedbound (8.9%), suggesting representation across all stages of disease.

### Symptoms of nOH and diagnostic certainty

The pre-specified critical dataset with the highest certainty of MSA with nOH included 108 respondents (out of 259 total MSA, 41.7%) with a physician diagnosis and ≥1 or more symptom upright, not present while supine. Nearly half of patients with MSA and nOH (49.5%) waited one year or more after first noticing symptoms of nOH before eventually being diagnosed. The most common symptoms of nOH included dizziness or lightheadedness (88.0%), fatigue or tiredness (79.6%), weakness (76.9%), head/neck pain (50.9%) and visual problems (49.1%). Poor concentration and unresponsiveness occurred in 38.0% and 29.6%, respectively. Less common symptoms included orthostatic dyspnea (25.9%) and chest pressure/pain (5.6%). The vast majority reported symptoms standing (87.0%), arising from a chair (75.9%), or walking (57.4%), while a small but significant proportion endorsed the presence of symptoms while seated (15.7%). Symptoms were most noticeable in the morning (42.6%) or after meals (consistent with post-prandial hypotension, 25.0%). Nearly a quarter of MSA patients with nOH reported frequent fainting when standing or walking (24%)

To investigate the possibility of underdiagnosis of nOH in patients with MSA, we identified a subgroup of 103 respondents (out of 259 total MSA, 40.0%) without a diagnosis of nOH, but one or more classic nOH symptoms occurring in the upright position, that interfered with the activities of daily living. The frequency and prevalence of other autonomic symptoms in this “MSA with suspicion of nOH” group—including gastrointestinal dysfunction (70.9%), urinary retention (40.8%), nocturia (48.5%) and sudomotor impairment (46.6%)—was similar compared to those with a diagnosis of nOH, suggesting the overlapping of widespread autonomic failure in both groups (see Table 1). Remarkably, 21.2% of MSA patients reported having never had their BP measured in the upright position.

### Associations between nOH symptoms and daily functional burden

Table 2 shows the activities of daily living that were most limited by nOH. In MSA with nOH patients, daily activities that combined the upright position with heat stress, mild exercise, or sudden postural change were most affected. Half (49.5%) of patients reported planning their day around their symptoms, indicative of an adaptive behavioral component. In total, 75.7% reported needing help from their caregivers for symptoms related to nOH. In MSA patients with nOH and suspicion of nOH, we observed a positive, albeit moderate, relationship between the number of reported nOH symptoms and the number of activities of daily living that were limited by BP drops, *r*(198)=0.47 95% CI [0.35, 0.57], *p*=2.80 x 10^−12^ (see Figure 2A).

**Figure 2.**
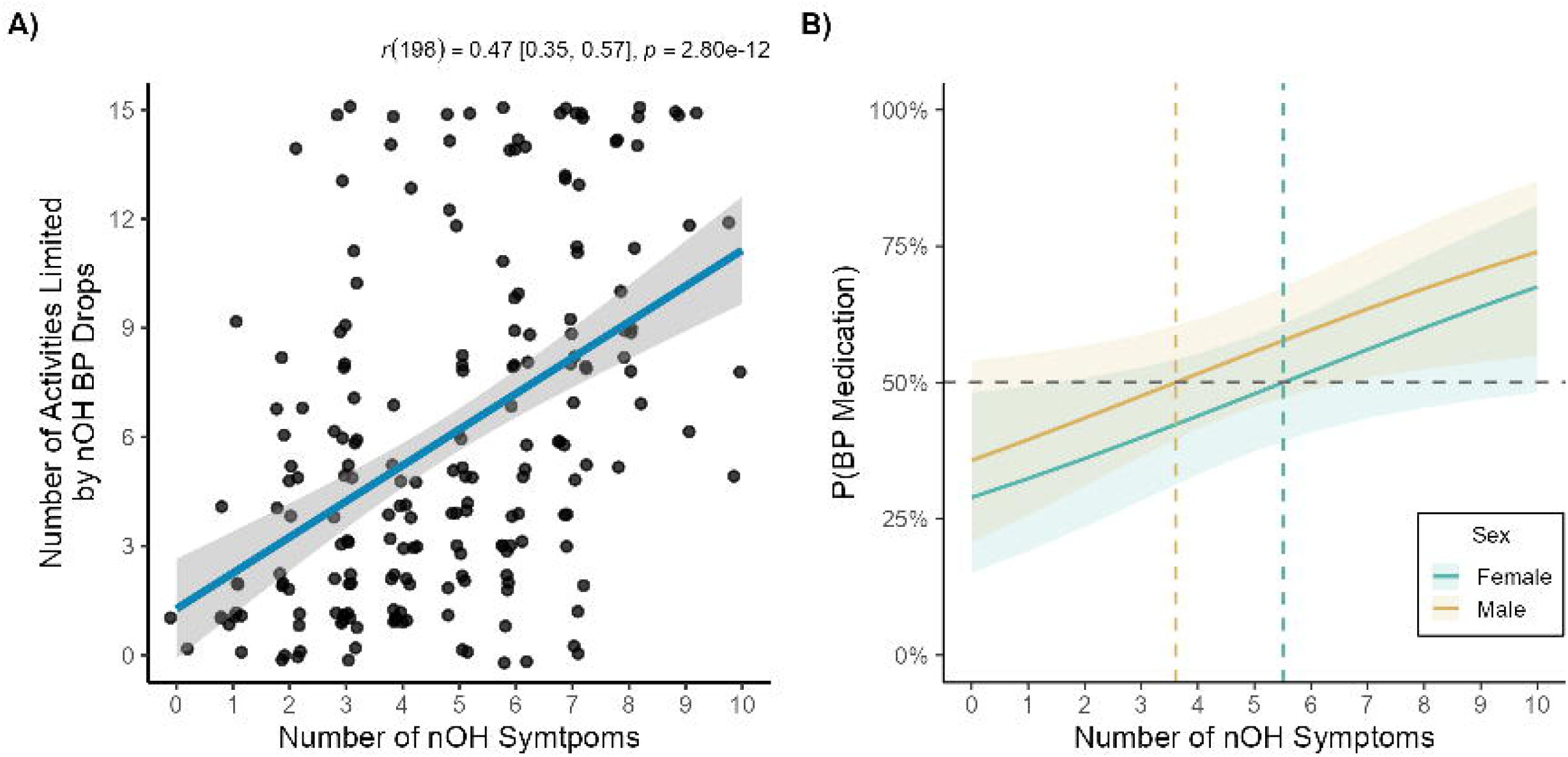
In MSA participants with nOH and with suspicion of nOH, the number of reported nOH symptoms were positively associated with A) functional burden and B) the probability of taking blood pressure (BP) medications. Shading denotes 95% confidence intervals.

**Table 2.** Neurogenic orthostatic hypotension symptom burden, treatment, and aspirations in the MSA cohort and diagnostic certainty subgroups.

| Parameter | Full MSA Population | MSA with nOH | MSA with Suspicion of nOH |
| --- | --- | --- | --- |
| n (% of Full MSA Population) | 259 (100%) | 108 (41.7%) | 103 (39.77%) |
| Total Number of nOH Symptoms Endorsed | 4.67 (0.15) | 4.82 (0.21) | 4.79 (0.23) |
| **Positions/Characteristics Associated with Symptoms** |  |  |  |
| Standing | 168 (68.29%) | 94 (87.04%) | 73 (70.87%) |
| Walking | 115 (46.75%) | 62 (57.41%) | 52 (50.49%) |
| Sitting | 45 (18.29%) | 17 (15.74%) | 28 (27.18%) |
| Supine | 23 (9.35%) | 0 (0%) | 21 (20.39%) |
| Overwhelming Urge to Sit/Lie When Standing | 98 (38.58%) | 42 (38.89%) | 45 (43.69%) |
| Need for Help from Family or Caregivers | 120 (61.54%) | 81 (75.7%) | 38 (43.68%) |
| Planning Day Around Symptoms | 77 (39.69%) | 53 (49.53%) | 24 (27.59%) |
| **Severity of nOH Symptoms** |  |  |  |
| Always Present/Affecting Most of the Day | 34 (16.35%) | 23 (21.3%) | 11 (11.11%) |
| Often Bothersome/Frequently Disruptive | 47 (22.6%) | 30 (27.78%) | 17 (17.17%) |
| Sometimes Bothersome/Could Get In Way | 47 (22.6%) | 23 (21.3%) | 23 (23.23%) |
| Occasionally Bothersome/Not Disruptive | 51 (24.52%) | 26 (24.07%) | 25 (25.25%) |
| Unnoticeable |  |  |  |
| **Activities Limited by nOH Symptoms** |  |  |  |
| Getting Out of Bed | 92 (45.77%) | 55 (50.93%) | 36 (39.13%) |
| Brushing Teeth | 51 (25.37%) | 30 (27.78%) | 21 (22.83%) |
| Getting Up from a Chair | 110 (54.73%) | 61 (56.48%) | 48 (52.17%) |
| Using the Toilet | 76 (37.81%) | 45 (41.67%) | 31 (33.7%) |
| Showering/Bathing | 109 (54.23%) | 65 (60.19%) | 43 (46.74%) |
| Climbing Stairs | 87 (43.28%) | 52 (48.15%) | 34 (36.96%) |
| Standing for a Short Time | 70 (34.83%) | 41 (37.96%) | 29 (31.52%) |
| Standing in a Line To Pay | 82 (40.8%) | 50 (46.3%) | 31 (33.7%) |
| Standing to Cook/Clean Dishes | 89 (44.28%) | 58 (53.7%) | 31 (33.7%) |
| Walking Between Rooms | 74 (36.82%) | 44 (40.74%) | 30 (32.61%) |
| Walking to End of Street | 74 (36.82%) | 48 (44.44%) | 26 (28.26%) |
| Taking a Longer Leisure Walk | 72 (35.82%) | 47 (43.52%) | 25 (27.17%) |
| Light Housework | 83 (41.29%) | 56 (51.85%) | 27 (29.35%) |
| Shopping for Food | 69 (34.33%) | 46 (42.59%) | 23 (25%) |
| Visiting Family/Friends Outside the Home | 66 (32.84%) | 41 (37.96%) | 25 (27.17%) |
| Any of the Above | 187 (93.03%) | 102 (94.44%) | 84 (91.3%) |
| Total Activities Limited | 5.99 (0.33) | 6.84 (0.46) | 5 (0.45) |
| **Pharmacological Treatment of nOH<sup>2</sup>** |  |  |  |
| Use of Non-Pharmacological/Lifestyle Measures<sup>1</sup> | 169 (87.11%) | 101 (94.39%) | 68 (78.16%) |
| Takes BP Medication(s) | 97 (50.26%) | 77 (71.96%) | 20 (23.26%) |
| Use of 2 or More Drugs | 46 (47.92%) | 39 (51.32%) | 7 (35%) |
| Considers Symptoms Well Controlled | 0 (0%) | 0 (0%) | 0 (0%) |
| **Aspirations for Better Treatments** |  |  |  |
| To Feel Safer Standing or Walking | 120 (64.86%) | 75 (73.53%) | 45 (54.22%) |
| To Change Position Without Symptoms | 95 (51.35%) | 60 (58.82%) | 35 (42.17%) |
| To Stand for Longer Durations | 121 (65.41%) | 74 (72.55%) | 47 (56.63%) |
| To Walk Longer Distances | 109 (58.92%) | 66 (64.71%) | 43 (51.81%) |
| To Be More Independent | 109 (58.92%) | 73 (71.57%) | 36 (43.37%) |
| To Reduce Family Worry | 98 (52.97%) | 69 (67.65%) | 29 (34.94%) |

| Parameter | Full MSA<br>Population | MSA with<br>nOH | MSA with<br>Suspicion of<br>nOH |
| --- | --- | --- | --- |
| To Socialize More | 100 (54.05%) | 63 (61.76%) | 37 (44.58%) |
| Shower or Bathe Without Fear of Fainting | 90 (48.65%) | 60 (58.82%) | 30 (36.14%) |
| Do Household Chores More Easily | 69 (37.3%) | 43 (42.16%) | 26 (31.33%) |

*Note.* Continuous parameters are presented Mean (SEM); categorical parameters are presented n (%); the 'n' row percentage represents each subgroup's proportion of the Full MSA Population; all other row percentages are calculated from valid (non-missing) responses within each group.
<sup>1</sup> Includes salt, water, head-up sleeping, abdominal binder, compression stockings, or smaller meals.
<sup>2</sup> Includes fludrocortisone, midodrine, droxidopa, pyridostigmine, atomoxetine, or other.

The MCA using a total of 31 symptoms—motor (4), cardiovascular autonomic (11), other autonomic (11), and cognitive/mood (5)—across 207 participants from dataset #2 (49% MSA with suspicion of nOH) resulted in four significant PCs via permutation testing (see Figure S1). By examining significant loadings via computed bootstrap ratios (see Figure 3) and geometrically plotted loadings (see Figure S2), we observed that PC1 explained 74% of the variance (*p*=0.0005) and captured general MSA symptom burden with all the symptoms loading onto PC1 in the same direction. PC2 explained 13% of the variance (*p*=0.0005) and contrasted urinary dysfunction (catheter use, urinary retention UTIs) with nOH symptoms. PC3 explained 4% of the variance (*p*=0.0225) and opposed signs of autonomic failure (hypohidrosis, catheter use) with cognitive/mood symptoms (anxiety, depression, nOH-related cognition). Using PC regression, we observed that only general MSA symptom burden (PC1) was associated with functional burden (see Table 3). Table S1 reports additional regression model information and sample sizes for all models.

**Figure 3.**
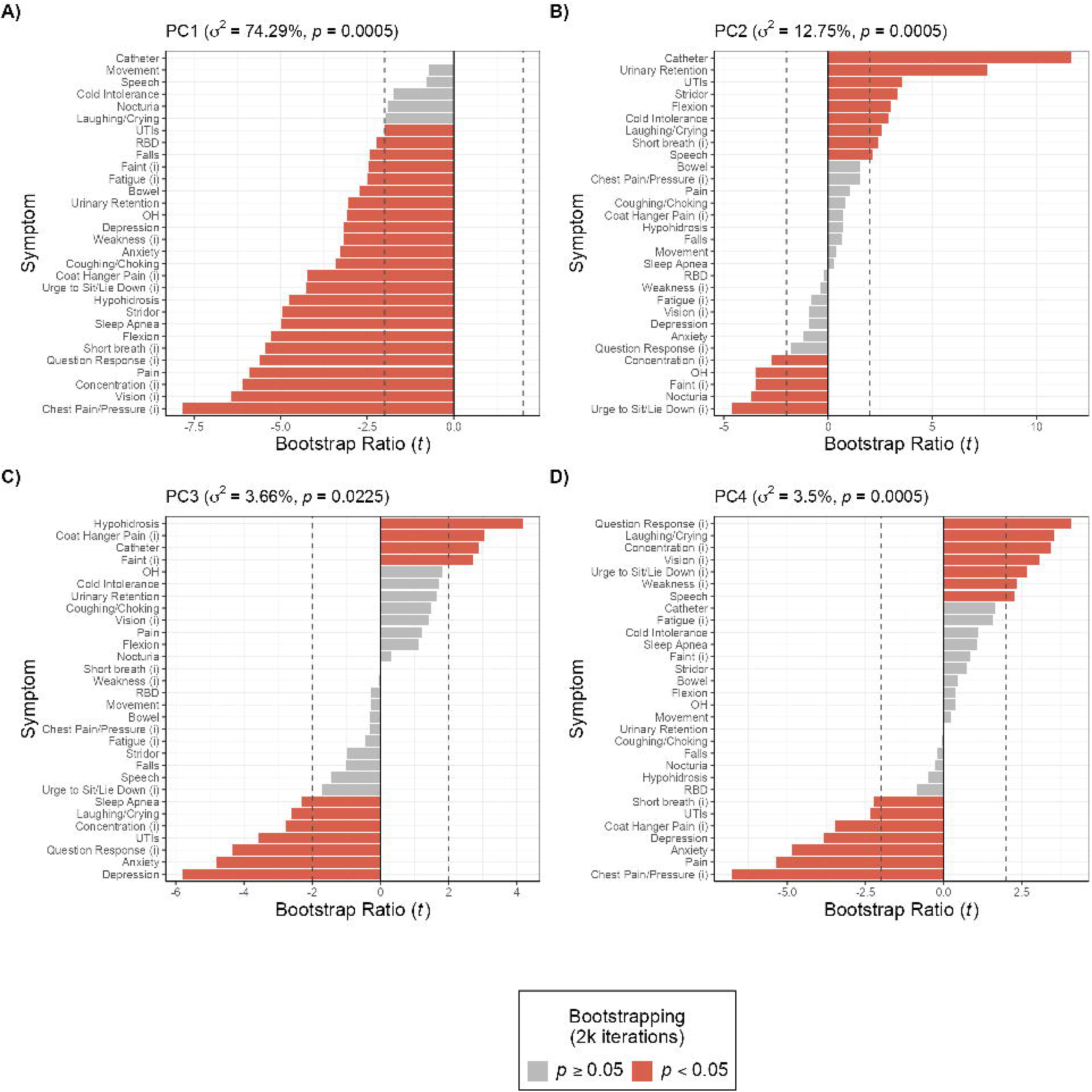
Symptom loadings reveal distinct dimensions of multiple system atrophy (MSA) and neurogenic orthostatic hypotension (nOH) symptomology. Bootstrap ratios >2 indicate symptoms that reliably/significantly contribute to the variance of each principal component (PC).

**Table 3.** Linear and logistic regression predicting neurogenic orthostatic hypotension functional burden and blood pressure medication use.

| **Linear Regression: Total Activities Limited by nOH** |  |  |  |  |  |
| --- | --- | --- | --- | --- | --- |
| **Predictor** | **b** | **95% CI** | **SE** | **t** | **p** |
| Intercept | 5.92 | [5.03, 6.81] | 0.45 | 13.12 | 1.84e-28 |
| Sex (Male) | 0.23 | [-1, 1.45] | 0.62 | 0.36 | 0.717 |
| Age | 0.20 | [-0.38, 0.79] | 0.30 | 0.69 | 0.494 |
| PC1: General MSA symptom burden | -2.33 | [-2.91, -1.74] | 0.30 | -7.87 | 2.52e-13 |
| PC2: Urinary symptoms with fewer nOH symptoms | -0.03 | [-0.61, 0.56] | 0.30 | -0.09 | 0.928 |
| PC3: Autonomic failure with fewer cog./mood symptoms | 0.27 | [-0.32, 0.86] | 0.30 | 0.90 | 0.371 |
| PC4: nOH-related pseudobulbar affect with fewer pain/cog./mood symptoms | -0.02 | [-0.62, 0.58] | 0.30 | -0.06 | 0.954 |
| **Logistic Regression: Taking BP Medications** |  |  |  |  |  |
| **Predictor** | **OR** | **95% CI** | **SE** | **z** | **p** |
| Intercept | 0.97 | [0.61, 1.55] | 0.24 | -0.13 | 0.895 |
| Sex (Male) | 1.19 | [0.62, 2.3] | 0.33 | 0.52 | 0.600 |
| Age | 1.26 | [0.92, 1.73] | 0.16 | 1.44 | 0.149 |
| PC1: General MSA symptom burden | 0.61 | [0.43, 0.83] | 0.17 | -3.02 | 0.003 |
| PC2: Urinary symptoms with fewer nOH symptoms | 1.05 | [0.77, 1.43] | 0.16 | 0.28 | 0.779 |
| PC3: Autonomic failure with fewer cog./mood symptoms | 1.54 | [1.13, 2.14] | 0.16 | 2.64 | 0.008 |
| PC4: nOH-related pseudobulbar affect with fewer pain/cog./mood symptoms | 1.12 | [0.82, 1.55] | 0.16 | 0.72 | 0.470 |

Note. Linear regression estimates are unstandardized b coefficients; logistic regression estimates are odds ratios (OR). SE = standard error on the model's native scale (log-odds for logistic). 95% CI = confidence interval. PC = principal component; all continuous predictors are standardized (M = 0, SD = 1). Sex reference category = Female.

### Treatment patterns and component interactions

In MSA patients with nOH, 94.4% were using non-pharmacological interventions, 72.0% reported taking medications for OH, and 51.3% were treated with two or more anti-hypotensive medications. The most common prescribed drugs were midodrine (73.7%), fludrocortisone (46.1%), pyridostigmine (23.7%), droxidopa (26.3%) and atomoxetine (9.2%). Anti-hypotensive medications were most often prescribed by a neurologist (53.3%), general practitioner (17.3%) or cardiologist (12.0%).

Maximizing the Youden index revealed that 6 nOH symptoms was the optimal cut point for BP medication use; however, given the poor classification performance (AUC=0.583, sensitivity=0.469, specificity=0.659) it is unlikely that nOH symptom burden alone classified BP medication use. With logistic regression, we observed that each additional reported nOH symptom resulted in 18% greater odds of taking BP medication (*OR*=1.18 [1.03, 1.36], *p*=0.021), adjusted for age (years: *OR*=1.03 [0.99, 1.07], *p*=0.163) and sex (males: *OR*=1.36 [0.76, 2.48], *p*=0.31). While holding age constant at the sample mean (*M*=64.66, *SD*=8.16 years) predicted probability curves for males and females resulted in between 3-6 symptoms, respectively, as the threshold at which the probability of BP medication use was 50% (see Figure 2B).

Using PC regression, we observed that hypohidrosis (PC3) was associated with greater odds of BP medication use in addition to, and independent of, general MSA symptom burden (PC1; see Table 3). Including MSA disease duration (in years) as a covariate in all linear and logistic models did not change the results. Given that nearly 70% of MSA participants received a different initial diagnosis and visited four physicians on average prior to an MSA diagnosis, MSA disease duration may not be accurately captured; therefore, we present the more parsimonious model estimates herein.

### Unmet needs

Finally, we wanted to determine the impact of treatment on nOH symptoms. Of the patients receiving anti-hypotensive drug treatment, none considered their symptoms well controlled, and refractory symptoms interfered with one or more daily functions in 97.1% of respondents. The most notable remaining limitations included difficulty with kitchen chores, showering/bathing, arising from a chair, light housework, getting out of bed, and climbing the stairs. MSA patients with nOH reported that they want better management of their low BP to feel less lightheaded or dizzy (80.8%), less fatigued (68.3%) and less weakness (68.3%). They reported wanting to feel safer when standing or walking (73.5%), stand for longer without needing to sit (72.5%), become more independent (71.6%), and for their family to worry less about their symptoms of nOH (67.6%). The presence of supine hypertension resulted in patients having to stop, reduce, switch OH medications or add additional antihypertensive medications in 29.1% of cases.

## Discussion

In this community-based cross-sectional study involving 259 patients with MSA, we show that nOH is likely to be underdiagnosed and undertreated. Over 40% of MSA patients have symptoms consistent with autonomic failure and organ hypoperfusion on standing, but no diagnosis of nOH. These data show that the odds of being diagnosed with nOH and receiving treatment with an anti-hypotensive drug are highest in the subgroup of patients with more severe autonomic failure, extending to the bladder and sudomotor function. Despite treatment with anti-hypotensive drugs, over 97% of patients with nOH remain symptomatic and functionally limited by nOH. In this real-world dataset, we found that patients with three or more symptoms of nOH had a predicted probability of 50% of taking BP medications, but the true functional impact of nOH is heavily influenced by background disease.

These data show patients wait on average 3 years from first symptom onset to MSA diagnosis and have a 70% chance of being first misdiagnosed, which is a call to action. Despite efforts to revise diagnostic guidelines and include “prodromal MSA” [28], earlier diagnosis has not translated into patient experience data, with the delay remaining substantial and unchanged for over a decade [29]. Even in the absence of disease-modifying therapies, timely diagnosis is still important as it enables early symptomatic interventions that help maintain mobility [30]. What emerges from the data is a high frequency of healthcare use prior to diagnosis, which is a signal known to precede the onset of a rare disease [31]. The question remains whether there are signature healthcare utilization patterns in the early stages of MSA that could translate into practical actionable criteria to support timely diagnosis and prompt care.

These data capture, directly from patients, the current gaps in knowledge around nOH in MSA. First, they point to the clear lack of awareness of autonomic failure within the medical community, with 21% of MSA patients reporting having *never* had their BP measured in the standing position at a clinical visit. Awareness of supine hypertension, which often coincides with nOH [32], was equally lacking. Second, at least 40% of MSA patients have recognizable and burdensome symptoms of global cerebral ischemia when standing [33], but no diagnosis of nOH. These results indicate the high likelihood that nOH is underdiagnosed and missed at a community level because orthostatic blood pressures are not routinely taken.

A novel aspect of our study is the use of MCA, a variant of principal component analysis, on patient reported data to understand the complex interactions background disease status and the true functional impact of nOH. Logistic regression identified a relationship between the number of symptoms reported and need for treatment with anti-hypotensive drugs. Using MCA, we expanded on this to show that BP drug treatment is associated with hypohidrosis and catheter use independent of general MSA symptoms. This suggests the presence of an identifiable subgroup of patients with more severe widespread autonomic failure associated with the treatment with anti-hypotensive drugs.

A major goal of our study was to determine the functional impact of nOH on the lives of patients with MSA. We show that despite drug treatment, not one MSA patient considers their symptoms well-controlled, which is thought provoking. There are only two approved pressor agents and a handful of off-label drugs used in the treatment of nOH, in contrast to over 100 drugs available for the management of hypertension, with many reformulated to allow once-daily dosing [34]. Refractory nOH symptoms impose limitations on MSA patients’ abilities to cook, bathe, and arise from the chair or bed, which cover all the basic skills required to maintain independence [35].

However, these data show that the functional burden of nOH is only moderately related to the number of symptoms of nOH, with the background disease accounting for substantial variance. This has implications for assessing nOH symptom burden with subjective measures. We show that the complexity of the disease adds noise to the interpretability of symptom measures, which will need to be overcome with statistical methods for a treatment effect to emerge in a clinical trial. Currently, the only phase III symptomatic endpoint in clinical trial readouts is the subjective OHQ, which relies on past week symptom recall at one time point, is heavily impacted by intercurrent events (like falls, fractures, infections), periods of immobility, and by mood. This makes it difficult to replicate findings from trial-to-trial. The need to develop sensitive objective endpoints to quantify the functional burden of nOH remains paramount.

A major advantage of our study is the community-based online survey approach, which allowed us to achieve in months what would have taken years to achieve with conventional clinical-based research visits. This approach improves representation from MSA patients living with advanced stage disease or in remote areas who are often underserved [36]. The prevalence of both motor and non-motor signs and symptoms aligns with clinic-based prospective natural history cohort studies of MSA in the Europe and North America [13, 37], suggesting this was a representative cross-sectional sample. To the best of our knowledge, this is the largest online research survey in MSA focused on nOH [29, 36], conducted in under six months, and a compelling argument of the power of this approach in rare diseases.

Our survey went through a rigorous development process but has some limitations. Like similar survey studies, the diagnosis of MSA is based on self-report and required web access. Although there are studies in Parkinson disease that corroborate the high degree of accuracy between a self-reported and clinician-made diagnoses [38], no such study exists yet for MSA. Nevertheless, a high proportion of participants received their MSA diagnosis from a neurologist, which adds confidence. We applied an age cut off (<80 years), which may have excluded some late onset MSA cases [39]. The survey was developed to obtain the patient’s (not caregiver) experience. As a result, non-response bias cannot be ruled out, but advanced stage disease patients are represented within the cohort. The survey was anonymized and we did not collect geographic information, so we cannot examine the experiences of patients in different regions nor identify/exclude duplicate submissions. We did not include a comparative cohort with other synucleinopathies, which would have been interesting, but beyond the scope of the present study.

This study found that nOH is underrecognized and undertreated in patients with MSA, leading to substantial functional limitations, loss of independence, and caregiver burden. While awareness of nOH and access to care remain major hurdles, it is worthwhile noting that 91% of MSA patients have access to a BP monitor at home, which is twice the national average [40]. This suggests the possibility of developing a tool to obtain real-world standardized BP measures to screen for nOH, which is more often detected on home readings than in a physician office [41]. It is our hope that these findings are leveraged to advocate better treatments for nOH in patients with MSA, which are urgently needed.

## Supporting information

CROSS checklist

## Data Availability

Data are available on Open Science Framework (link to be inserted here upon publication).

## Acknowledgement

We thank all the patients and their caregivers for giving up their valuable time to participate. We thank MSA Trust UK, Defeat MSA, and Mission MSA for their support.

## Author Roles

(1) Research project: A. Conception, B. Organization, C. Execution; (2) Statistical Analysis: A. Design, B. Execution, C. Review and Critique; (3) Manuscript Preparation: A. Writing of the first draft, B. Review and Critique;

MJK: 1B, 1C, 2ABC, 3AB

LO: 1ABC, 3B

MS: 1ABC, 3B

VI: 1AC, 3B

RF: 1A, 3B

JJ: 1A, 3B

IB: 1A, 3B

HK: 1A, 3B

TG: 1A, 2C

RV: 1A, 3B

AM: 1A, 3B

ES: 1BC, 3B

ER: 1BC, 3B

LV: 1AC, 2B, 3B

LJNK: 1ABC, 2AC, 3AB

## Ethical Compliance Statement

The study protocol and recruitment materials were reviewed by two external ethics committees (US: Advarra and UK: Health Research Authority Research Ethics Committee), and exemptions granted in both countries. Patient consent was obtained online via a web-based consent process (see Supplemental Material). We confirm that we have read the Journal’s position on issues involved in ethical publication and affirm that this work is consistent with those guidelines.

