## Supplementary Material for "The burden of neurogenic orthostatic hypotension in patients with multiple system atrophy – a real-world study"

Last modified: 2026-04-20 18:18:10

#### Table of contents

**1 Survey** **5**
- 1.1 Survey Questions . . . 5
- 1.2 Resource Information for Patients (extracts) . . . 10

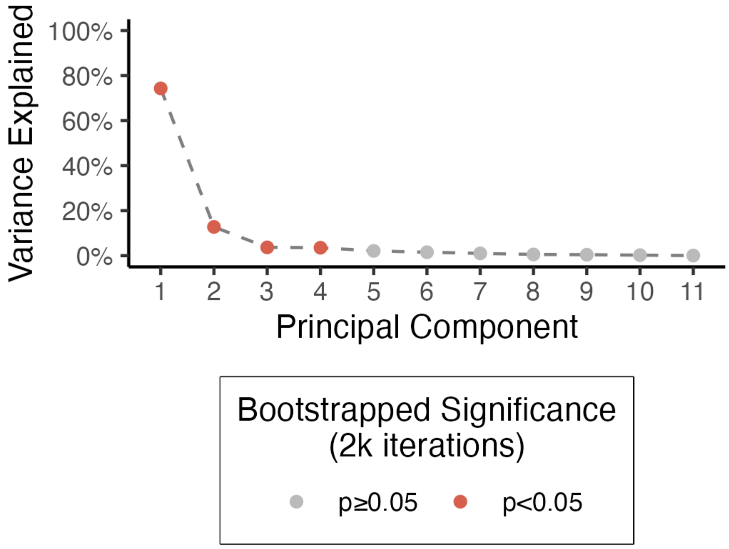

Figure 1: Scree plot showing variance explained per component. Red points indicate components significant at $p<0.05$ via permutation testing (2k iterations).

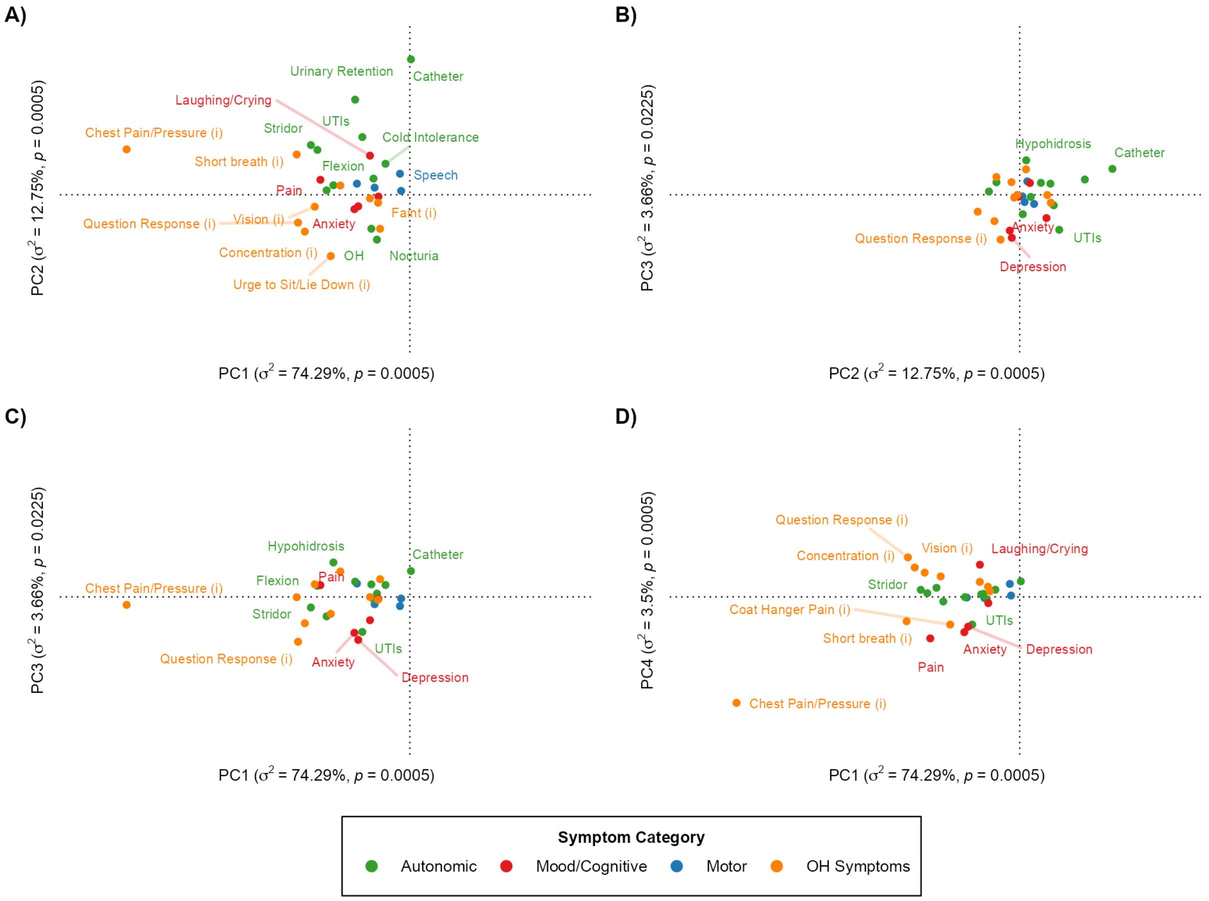

Figure 2: Column factor score plots for principal components (PCs) 1–4 displayed in pairs. Each point represents the symptom-present category of a binary symptom indicator; its position reflects how strongly that symptom contributes to each component, with symptoms closer together tending to co-occur among respondents. Point color indicates symptom domain (Motor, Autonomic, Mood/Cognitive, OH Symptoms). Labels marked '(i)' denote symptoms experienced while standing or walking that improve upon sitting or lying down. Axis scales are held constant across all panels to allow direct comparison of relative positions.

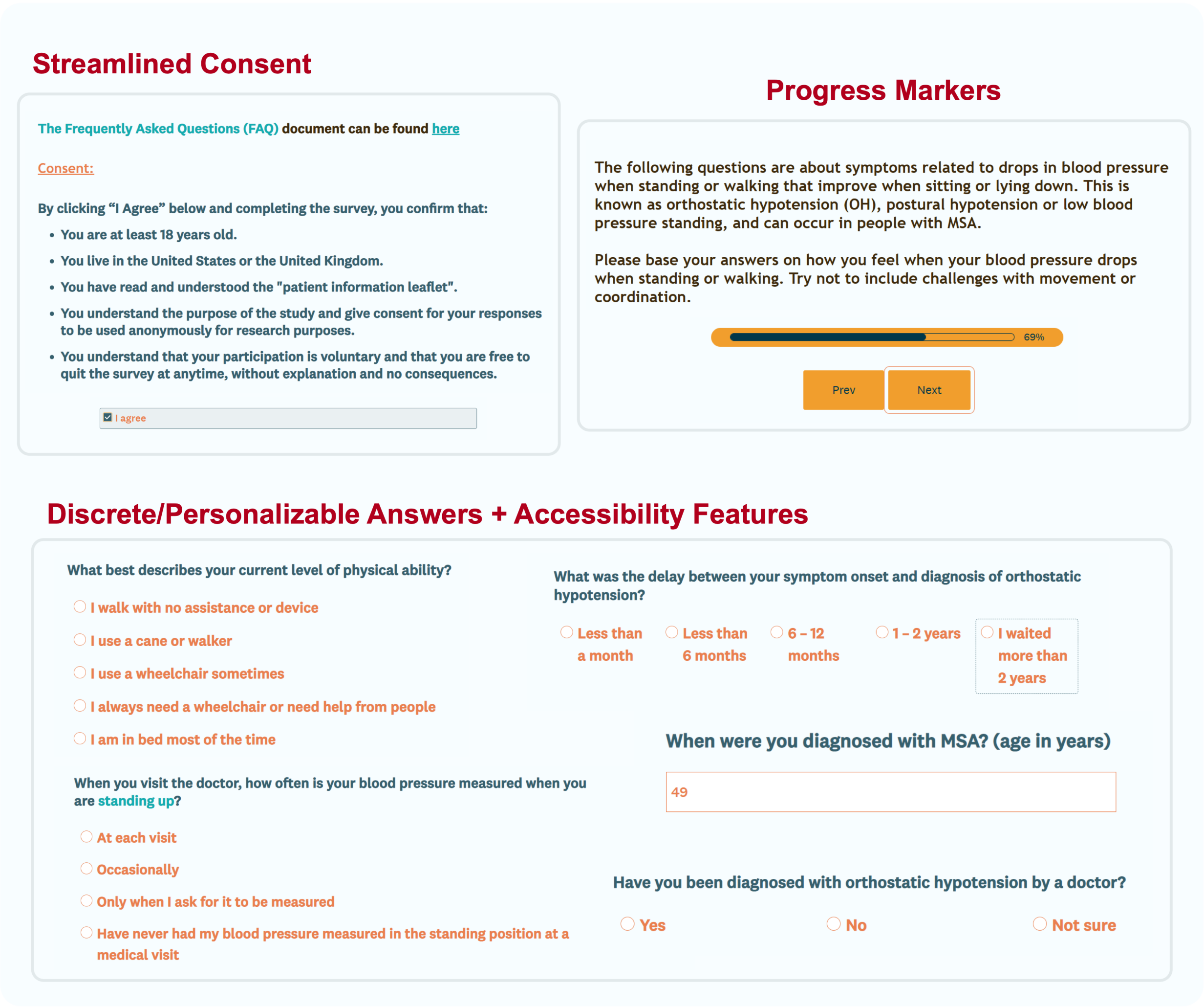

Figure 3: Web-based participant facing data collection platform for the 'Living with MSA: The Low Blood Pressure Symptom Survey'.

Table 1: Regression model details: formulas, sample sizes, and model fits.

| Model | $N$ (% nOH[a]) | $R^2$ | Model Fit | $p$ |
|---|---|---|---|---|
| Functional Burden ∼ PC1–4 + Sex + Age (linear) | 198 (53.5%) | 0.249 | $F(6, 191) = 10.55$ | $4.16 \times 10^{-10}$ |
| BP Medication Use ∼ PC1–4 + Sex + Age (logistic) | 187 (56.1%) | 0.078 | $\chi^2(6) = 20.19$ | 0.003 |
| BP Medication Use ∼ No. nOH Symptoms + Sex + Age (logistic) | 187 (56.1%) | 0.031 | $\chi^2(3) = 7.93$ | 0.047 |

*Note.* MSA=multiple system atrophy; nOH=neurogenic orthostatic hypotension; PC=principal component; BP=blood pressure. $R^2$ is the coefficient of determination for the linear model and McFadden's pseudo-$R^2$ for logistic models.
[a] Percentage of participants classified as MSA with nOH (highest certainty); remaining participants were classified as MSA with suspicion of nOH.

### 1 Survey

The "Living with MSA: The Low Blood Pressure Symptom Survey" was hosted on SurveyMonkey and administered by Klick Health. Theravance parterned with Klick to run the survey. Klick is a specialist health research company with extensive experience in running patient surveys.

Participants were adults ($\geq$ 18 years) living with MSA in the United States or United Kingdom. The survey comprised 28 items organized into two parts: Part 1 covering MSA diagnosis history and symptom burden, and Part 2 covering orthostatic hypotension symptoms, functional impact, management, and treatment. Screenshots of the survey platform are provided in Figure 3.

#### 1.1 Survey Questions

##### Introduction and Consent

Participants were presented with a title banner, a recruitment graphic, and an educational infographic about orthostatic hypotension. They then read an "About our survey" page and a consent section confirming eligibility (age $\geq$ 18 years, have MSA, residing in the US or UK) and voluntary participation before proceeding.

##### Part 1: MSA Diagnosis and Symptoms

**Q1.** Do you have MSA? (Yes / No / Not sure)

**Q2.** What is your current age? (open numeric, in years)

**Q3.** What is your biological sex? (Male / Female / Other)

**Q4.** When did your symptoms of MSA first start? (age in years)

**Q5.** When were you diagnosed with MSA? (age in years)

**Q6.** Before your diagnosis, how many different doctors did you visit for symptoms that were later found to be related to MSA? (open numeric)

**Q7.** Were you first told you had a different diagnosis than MSA? (Yes / No / Not sure)

- *If yes → Q7A:* What was your first diagnosis? (Parkinson's disease / Dysautonomia or pure autonomic failure / A psychological disorder / Cerebellar ataxia / Atypical Parkinson's or Parkinson's plus syndrome / Other / Not sure)

**Q8.** Who first diagnosed you with MSA? (A neurologist / An autonomic nervous system specialist / A cardiologist / A sleep medicine doctor / A urologist / Primary care/general practitioner / Other / Not sure)

- *If neurologist → Q8A:* Was this a neurologist specializing in movement disorders or Parkinson's disease? (Yes / No / Not sure)

**Q9.** What symptoms of MSA do you currently have? (tick all that apply)

- *Motor and speech:* Problems with movement (clumsiness, tremor or jerking) / Falls / Coughing or choking / Slurred or soft speech
- *Non-motor:* Feeling dizzy/lightheaded/faint on standing / Inability to pee / Waking up more than once at night to pee / Frequent UTIs / Need for urinary catheter / Constipation or bowel accidents / Cold hands and/or feet / Sweating less / High pitched or noisy breathing (stridor) / Pauses in breathing while sleeping (sleep apnea) / Abnormal bending of neck or spine
- *Behaviors and mood:* Acting out dreams / Uncontrollable laughing or crying / Pain / Depression / Anxiety / Other

**Q10.** What best describes your current level of physical ability? (I walk with no assistance or device / I use a cane or walker / I use a wheelchair sometimes / I always need a wheelchair or need help from people / I am in bed most of the time)

##### Part 2: Orthostatic Hypotension Symptoms and Management

Participants were instructed to base answers on symptoms related to blood pressure drops when standing or walking, and to exclude challenges with movement or coordination.

**Q16.** Do you frequently faint or pass out when standing or walking? (Yes / No / Not sure)

**Q11.** Do you ever experience any of the following symptoms on standing or walking that improve when sitting or lying down? (check all that apply): Dizziness or lightheadedness or feeling like you might black out or faint / Difficulties with vision (blurred, darkened, or tunneled) / Pain across the neck and/or shoulders / Fatigue or tiredness / Weakness / Difficulty concentrating or paying attention / An overwhelming urge to sit or lie down / Difficulty responding to questions / Chest pain or pressure / Shortness of breath / Other / No, I do not experience any of these symptoms

- *If any symptom endorsed → Q11A:* When do you notice these symptoms? (check all that apply): When standing / When walking / When sitting / When getting up from a chair / When lying down / In all positions
- *If any symptom endorsed → Q11B:* At what time(s) of day do you notice your symptoms? (check all that apply): In the morning / Afternoon / Evening / Night / Before meals / After meals / Any time, with no clear pattern

**Q12.** On a typical day, approximately how long are you able to stand for without feeling these symptoms? (Less than a minute / 1–5 minutes / 5–10 minutes / For as long as I need / I can no longer stand or walk / Not sure)

**Q13.** Looking back over the past week, how would you rate your symptoms related to your blood pressure dropping while standing or walking? (I didn't notice them / I noticed them occasionally, but they did not affect my activities / They bothered me sometimes and could get in the way / They bothered me often and frequently disrupted my activities / They occurred each time I stood and affected most of my day)

**Q14.** Do these symptoms related to your blood pressure dropping on standing or walking limit any of your abilities to do the following activities? (tick all that apply): Getting out of bed / Brushing your teeth / Getting up from a chair / Using the toilet / Showering or bathing / Climbing the stairs / Standing for a short time / Standing in a line to pay / Standing to cook, wash dishes, or clean / Walking from one room to another / Walking to the end of a street / Taking a longer leisure walk / Doing light household chores / Shopping for food / Leaving the house to visit friends or family / Other

**Q15.** In your own words, which symptom that you feel when your blood pressure drops most limits your activities? (open text)

**Q17.** Have you been diagnosed with orthostatic hypotension by a doctor? (Yes / No / Not sure)

- *If yes → Q17A:* What type of doctor first diagnosed you with orthostatic hypotension? (A neurologist / An autonomic specialist / A cardiologist / A sleep medicine doctor / A urologist / Primary care / Other / Not sure)
  - *If neurologist → Q17B:* Was this a neurologist specializing in movement disorders or Parkinson's disease? (Yes / No / Not sure)
- *If yes or not sure → Q17C:* What was the delay between your symptom onset and diagnosis of orthostatic hypotension? (Less than a month / Less than 6 months / 6–12 months / 1–2 years / More than 2 years)

**Q18.** Do you need help from your family or caregivers due to your symptoms related to your blood pressure drops on standing? (Yes / No / Not sure)

**Q19.** Have you made any of the following lifestyle changes to help manage your symptoms related to your blood pressure dropping on standing? (check all that apply): Planning your day around your symptoms / Drinking extra water / Taking extra salt / Sleeping with the head of the bed raised / Wearing compression stockings / Using an abdominal binder / Eating smaller meals / Other / None

**Q20.** Do you take medications to help prevent your blood pressure drops on standing or walking? (Yes / No / Not sure)

- *If yes → Q20A:* What medications have you taken for your orthostatic hypotension? (Fludrocortisone / Midodrine / Droxidopa / Pyridostigmine / Atomoxetine / Other / Not sure)
- *If yes or not sure → Q20B:* Who writes the prescriptions for the medications you take to treat your blood pressure at follow-up? (A neurologist / A specialist nurse/nurse practitioner / An autonomic specialist / A cardiologist / A general practitioner/primary care doctor / Other / Not sure)

**Q21.** Do you have high blood pressure when lying down (also called supine hypertension)? (Yes / No / Not sure)

**Q22.** Have you had to make any of the following changes due to high blood pressure when lying down? (check all that apply): Add new medication(s) / Lower the dose of medication(s) / Switch to different medication(s) / Stop taking medication(s) / None of the above / Not sure

**Q23.** If you could better manage drops in blood pressure on standing, what symptoms would you most hope to see improve? (check all that apply): Less dizziness or lightheadedness / See more clearly / Think more clearly / Less fatigue / Less weakness / Less pain across shoulders / None (symptoms well controlled) / Other

**Q24.** If you could reduce your symptoms of blood pressure dropping on standing, what would you most wish to see? (check all that apply): Stand longer without needing to sit / Less difficulty changing positions / Walk longer without needing to rest / Feel safer when standing or walking / Do household chores more easily / Shower or bathe without fear of fainting / Spend more time outside the home / Socialize without worrying about symptoms / Family worrying less / Do more things independently / None (symptoms well controlled) / Other

**Q25.** Do you feel that your doctors are knowledgeable about blood pressure issues in people with MSA? (Yes / No / Not sure)

**Q26.** Do you have a blood pressure monitor or blood pressure cuff at home? (Yes / No / Not sure)

**Q27.** When you visit the doctor, how often is your blood pressure measured when you are standing up? (At each visit / Occasionally / Only when I ask / Never)

**Q28.** Finally, who is filling out this survey? (Completed by the patient / Completed by the caregiver / Completed by the patient and caregiver in equal proportion)

#### 1.2 Resource Information for Patients (extracts)

### PATIENT INFORMATION LEAFLET

#### LIVING WITH MSA: Low blood pressure symptoms

##### » What is MSA?

MSA is an abbreviation formed from the initial letters of the disease Multiple System Atrophy. MSA is a rare neurological disease that affects the brain and spinal cord. It impacts the ability of a person to move resulting in poor balance and coordination. It also impacts a person's ability to control the involuntary (autonomic) functions of the body including the bladder, bowel, blood pressure, heart rate, and or ability to sweat. People with MSA may also experience difficulty with swallowing, speaking, disrupted sleep, or pain. MSA used to be known as Shy Drager Syndrome, a term that is sometimes still used to describe the disease.

##### » What is orthostatic hypotension?

Orthostatic hypotension is the medical term for a drop in blood pressure in upright position. In people with MSA this is caused by problems with the nerves that should normally control blood pressure when standing or walking. If blood pressure drops when upright, it can fall to low levels, which reduces blood supply to the organs, and may cause symptoms. These symptoms can vary throughout the day and will improve when sitting or lying down. The diagnosis requires having blood pressure measurements taken when lying and standing during a doctor's visit. In some cases, people may be referred to an autonomic or cardiology clinic for a tilt-table test. Patients with MSA who have orthostatic hypotension may also experience high blood pressure in when lying down (a condition known as supine hypertension).

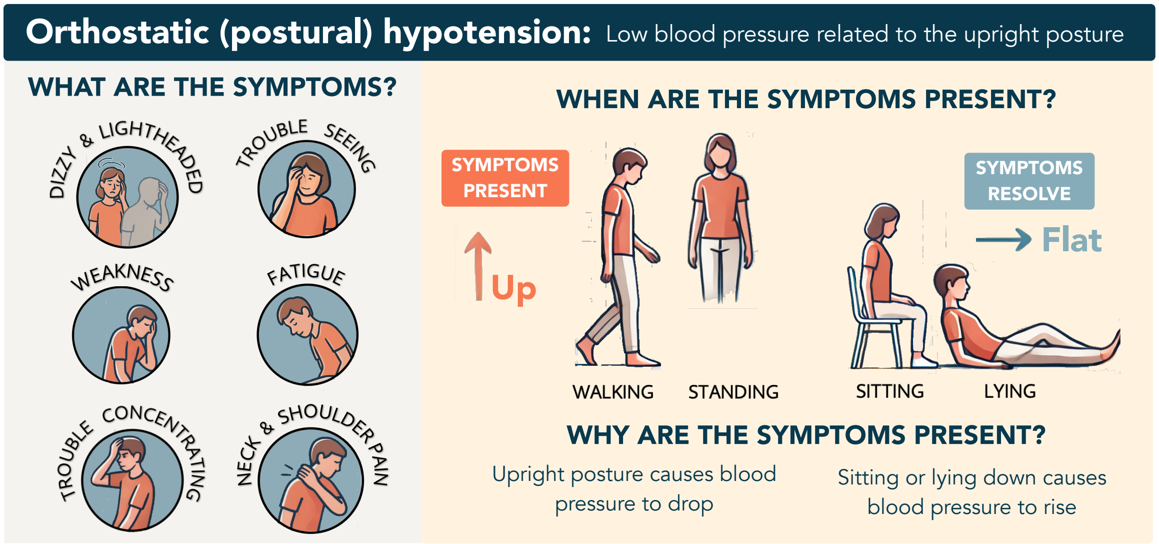

##### » What are drops in blood pressure in people with MSA?

Many people living with MSA experience drops in blood pressure when in the upright position. This condition goes by many names including **orthostatic hypotension**, postural hypotension, neurogenic orthostatic hypotension, dysautonomia, or autonomic failure. In people living with MSA, orthostatic hypotension is caused by a problem with the nerves that control blood pressure. Upon standing, blood pressure drops, often to low levels. Falls in blood pressure can lead to a reduced supply of blood to the organs, which can cause symptoms such as dizziness, lightheadedness, feeling faint, difficulty seeing, weakness, fatigue, pain across the back, and difficulty thinking or responding. These symptoms can vary throughout the day. They may be worsened with even slight exertion (e.g., climbing the stairs), in a warm room, or after meals. They improve when sitting or lying down.

#### » How can I tell if my symptoms are due to low blood pressure?

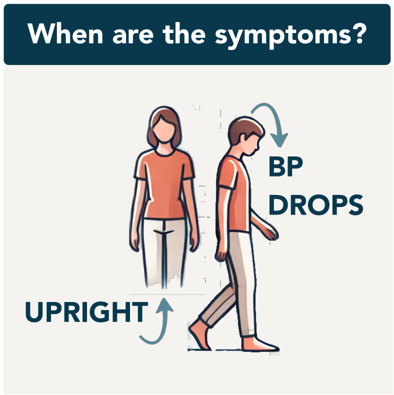

It may be challenging for people with MSA to tell when their symptoms are due to low blood pressure or are related to their problems with movement, coordination or imbalance. In general, symptoms of standing low blood pressure should have a clear association with changes in body position. They are often triggered when shifting to an upright position (e.g., going from lying to standing or sitting to standing). They may worsen after standing for some time, when walking, or with mild exertion. They improve when you sit and should stop when lying down. When upright, low blood pressure produces the sensation of fainting or near-fainting - rather than wobbliness, spinning or difficultly focusing due to uncontrollable eye movements.

#### » How will I know if I have been diagnosed with orthostatic hypotension?

- Orthostatic = related to or caused by the upright position
- Hypotension = low blood pressure

The diagnosis of orthostatic hypotension requires having blood pressure measurements taken when lying and standing during a doctor's visit. In some cases, people may be referred to an autonomic or cardiology clinic for a tilt-table test to diagnose their orthostatic hypotension. The diagnosis requires detecting a blood pressure drop of more than 20 mmHg systolic (top number) or 10 mmHg diastolic (bottom number) within 3 minutes of standing up. Occasionally, some clinics may diagnose orthostatic hypotension by checking blood pressure changes when shifting from a sitting to a standing position.

#### » How do I know if I am taking medications for orthostatic hypotension?

Your doctor may prescribe you medications that raise your blood pressure during the day. Depending on where you live, this may include midodrine (also called ProAmatine, Bramox or Gutron) or droxidopa (also called Northera). These drugs are known as anti-hypotensives (anti = against, hypotension = low blood pressure) or pressor agents. They are typically taken 2-3 times a day. You may have been told not to take these medications close to bedtime or skip doses when you are in bed, inactive, or lying down. Some people may also take a drug called fludrocortisone (also called Florinef). You may occasionally be prescribed other medications to treat your low blood pressure.

#### » What is supine hypertension?

Supine hypertension refers to high blood pressure when lying down in a person with orthostatic hypotension. This can occur in people with MSA and orthostatic hypotension. It is diagnosed when blood pressure is higher than 140/90 mmHg after 20 minutes lying flat and resting. Occasionally, people with supine hypertension may be prescribed medications for high blood pressure (known as anti-hypertensives) and or be told to sleep with the head of the bed raised. Supine hypertension is not usually associated with symptoms, but can be linked to increased blood pressure overnight and frequent need to urinate.

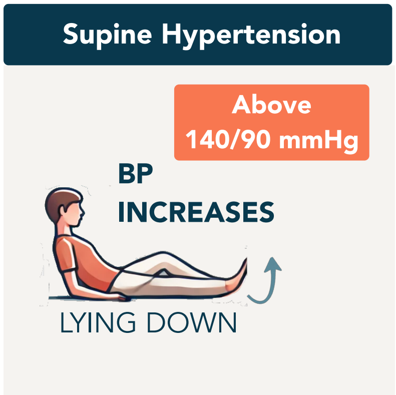
